# Prevalence and determinants of central obesity among adults 18–69 years in Kenya: a cross-sectional study

**DOI:** 10.1101/2024.09.18.24313881

**Authors:** Caleb Nyakundi, Sharonmercy Okemwa, Romeo Warera Ngesa, Samwel Maina Gatimu

## Abstract

**Background:** Central obesity affects about 4 out of every 10 people globally, and it is a notable public health concern linked with premature morbidity and mortality. In Kenya, regional studies have approximated the prevalence to range from 37% to 50%. However, these studies have been done in specific regions or population groups, such as among urban residents or women. Therefore, we aimed to assess the general and sex-specific prevalence and determinants of central obesity among adults in Kenya.

**Methods:** An analysis of the prevalence and determinants of central obesity was assessed using data from the 2015 Kenya STEPwise survey on non-communicable diseases and injuries. We included a final sample of 4 003 participants. Participants’ characteristics and prevalence of central obesity were described and summarized using frequencies and percentages. The bivariate and multivariate logistic regression were used to assess the determinants of central obesity.

**Results:** The overall weighted prevalence of central obesity was 46.7% [95% confidence interval (CI): 43.2, 50.3], with women exhibiting a significantly higher prevalence compared to men (58.3% vs. 35.6%, p=0.001). Sex, age, household wealth index, and smoking were among the factors significantly associated with central obesity. Men had 66% lower odds of central obesity compared to women, while the risk increased with age, ranging from 1.6 to 4 times higher odds among those aged 30—39 and 50—69 years, respectively, compared to 18—29–year–olds. Other predictors of central obesity were wealthier households and non-smoking.

**Conclusion:** About half of the adult Kenyan population is centrally obese, with a higher prevalence among women than men. Kenya’s policymakers should consider targeting high-risk groups in this population to reduce the burden of central obesity.

## Introduction

Central obesity is associated with an increased risk of metabolic syndrome, cardiometabolic dysregulation, and cardiovascular risk factors (1, 2, 3). Its economic cost is estimated at 2.2% of the global gross domestic product and is expected to rise to 3.3% by 2060 (4). In addition, it has a significant social impact often associated with stigma and discrimination, leading to social isolation, low self-esteem, and reduced quality of life (5, 6). The social burden of obesity extends to families and communities, as it can increase healthcare costs and reduce productivity (7). About four of every ten people are centrally obese worldwide (8). In sub-Saharan Africa (SSA), the prevalence of central obesity varies from country to country: 11.8% in Uganda (9), 40% in Ethiopia (10, 11), and 50.8% in Cote d’Ivoire (12). In Kenya, the prevalence of central obesity among adults ranges from 27.8% among undergraduate students (13) to 50% among adults living in informal settlements (14).

The risk of central obesity is linked to urbanization (15), sedentary lifestyle (16), dietary changes such as consumption of ultra-processed foods (17), and increased levels of physical inactivity (18, 19). In addition, studies have established that the prevalence of central obesity is higher among females, married individuals, urban residents, older adults, physically inactive individuals, and individuals with a certain level of formal education(20, 21, 22).

Kenya has committed to the Sustainable Development Goal (SDG) 3, target 3.4, which seeks to “reduce by one-third premature mortality from NCDs through prevention, treatment and promotion of well-being” (23). To reduce obesity rates, the Government of Kenya aims to reduce obesity rates from 27.9% to 25.8% by 2025 through minimization of exposure to modifiable risk factors and strengthening health promotion and education (24). To achieve these goals, there is a need for evidence on the magnitude and drivers of obesity.

Previous nationwide studies in Kenya on obesity have focussed on the prevalence and determinants of general obesity among adults, estimating it at 27% (38ꞏ5% of women and 17ꞏ5% of men) (25) (26), often overlooking central obesity. General obesity is characterized by abnormal or excessive fat accumulation across the entire body, but it differs from central obesity, which is characterized by the accumulation of fat in the abdominal region (27). While both forms of obesity are associated with increased health risks (28), central obesity is particularly detrimental due to its stronger association with cardiovascular diseases and other metabolic disorders (29, 30). However, few studies in Kenya (13, 14, 31) have focused on central obesity, with all of them not providing country-level estimates for the prevalence of central obesity. Therefore, this study uses the STEPwise survey data to address the paucity of evidence on the burden of central obesity and its determinants in Kenya. The study aims to assess the overall and sex-specific prevalence and determinants of central obesity in Kenya. The study results are key in contributing to the identification of appropriate interventions for the prevention and control of central obesity in Kenya.

## Methods

### Data source and study population

We conducted a secondary analysis using data from the 2015 Kenya STEPwise survey on risk factors for NCDs among adults aged 18 to 69 years (25, 32). This cross-sectional survey covered all 47 counties of Kenya, a lower-middle-income country with a population exceeding 54 million, with 70% residing in rural areas, 80% under the age of 35 and 38.6% living below the poverty line (33). The survey employed a three-stage cluster randomized approach. Initially, 200 clusters comprising rural and urban households were identified. Subsequently, 30 households were randomly selected from each cluster, and one individual was chosen from each household. Data collection followed the three steps of the World Health Organization (WHO) STEPwise approach, encompassing demographic and behavioural data collection (Step 1), anthropometric measurements (Step 2), and biochemical measurements of blood glucose and lipids (Step 3) (25). Of the 4,645 households included, 4,500 individuals participated in the survey (96.9% response rate).

### Measurements and variables

The primary outcome variable, central obesity, was assessed using waist circumference, hip circumference, and height measurements. Following the STEPwise approach (STEP 2), waist and hip circumferences were measured to the nearest 0.1 centimetres using a constant tape measure. The waist circumference was measured at the midpoint between the lowest rib and the hip bone at the end of a normal expiration with the arms relaxed at the sides, while the hip circumference was measured at the level of the greater trochanters. Participants who were pregnant, unable to stand, or had a colostomy or ileostomy were excluded from these measurements.

Prior to taking the measurements, participants were asked to remove any outer garments, such as jackets, bulky sweaters, cardigans, waistcoats, high-heeled shoes, and tight-fitting items like corsets, Lycra bodysuits, and support tights. If participants wore a belt, they were requested to remove or loosen it for the measurements.

A height/length measuring board was used to measure height in centimetres to the nearest millimetre. Participants were instructed to stand on the board without footwear, headgear, or elaborate hairstyles, with their feet together, heels against the backboard, and knees straight. Research Assistants received five days of training on conducting these measurements before the survey began.

#### Outcome variable

Central obesity was assessed using waist circumference (WC), waist-to-hip ratio (WHR), calculated by dividing waist circumference by hip circumference, and waist-to-height ratio (WHtR), obtained by dividing waist circumference by height. According to WHO criteria, individuals were classified as having central obesity and coded as “1” if they had a waist circumference of ≥80 cm for women or ≥94 cm for men, a WHtR >0.50, or a WHR of ≥0.85 for women or ≥0.90 for men (34); otherwise, they were coded as “0.“

#### Explanatory variables

Based on a review of the literature (13, 14, 18, 20, 21, 22) and with the availability of variables in the dataset, we selected and operationalized the demographic, socioeconomic and behavioural explanatory variables. The demographic variables included age, sex (male or female), locality of residence (urban or rural), and region of residence. Socioeconomic variables included the highest level of education, employment status, and household wealth quintiles generated by principal component analysis. The behavioural variables included smoking, alcohol consumption, fruit and vegetable intake and physical activity. Supplementary Table 1 describes and operationalizes the explanatory variables included in the study.

### Statistical analysis

We coded, cleaned, processed, and analyzed our data using STATA version 17 (35) and included participants without missing observations in the analysis (Fig. 1). Descriptive statistics were generated for the study variables using frequencies, proportions, and tables. To assess the association between central obesity prevalence among women and men and various respondent characteristics, we used Pearson’s chi-square test of independence. Also, we accounted for the survey’s cluster sampling design by applying the “svy” command in Stata and used the provided sample weights in the dataset to adjust for differing sampling proportions across enumeration areas.

**Fig 1.**
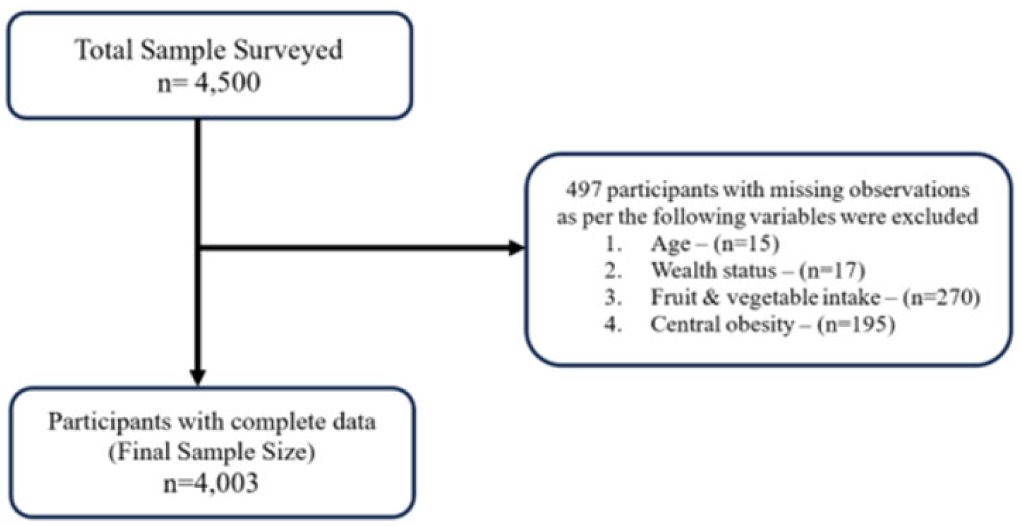
Flowchart of the study sample.

To identify factors associated with central obesity, we conducted both bivariate and multivariate binary logistic regression analyses. Prior to fitting variables into the multivariable model, we assessed multicollinearity among predictor variables using the variance inflation factor (VIF), setting a cutoff mean of 2.1, as recommended (36). Variables with p-values of less than 0.25 and clinically significant variables in the bi-variable analysis were considered and adjusted as possible predictors in the stepwise model selection for the multivariable logistic regression analysis model. The literature supports a cutoff value of 0.25, and also previous similar studies used the same cutoff (37, 38, 39). Finally, we calculated both crude and adjusted odds ratios with 95% confidence intervals to estimate the strength of the association between central obesity and participant characteristics, with statistical significance set at a p-value of <0.05.

### Ethics

The 2015 Stepwise survey participants provided written informed consent, and the Kenya Medical Research Institute Scientific Ethics and Review Unit approved the survey. The dataset was accessed from the Kenya National Bureau of Statistics data repository after a user’s agreement was completed and access was granted.

## Results

### Sample characteristics

Out of the 4500 participants in the survey, 4003 were included in the analysis after excluding 497 (11%) cases with missing observations to ensure consistency with the final sample (Fig. 1). Table 1 shows that most of the participants were: women (58.4%), married or in union (67.0%), self-employed (40.8%), and had no formal education (37.9%). More than three-quarters of the respondents participated in high-intensity physical activity (75.0%), had an inadequate intake of fruits and vegetables (75.8%) and had never consumed alcohol (78.1%) nor smoked (88.4%).

**Table 1.** Sample characteristics, prevalence, and factors associated with central obesity in Kenya (N = 4 003)

| Characteristics | Sample | Prevalence of central obesity |  | Bivariate logistic regression analysis | Multivariable logistic regression analysis |
| --- | --- | --- | --- | --- | --- |
|  | n (%) | n | % (95% CI) | cOR (95% CI) | aOR (95% CI) |
| **Sex** |  |  |  |  |  |
| Male | 1664 (41.6) | 633 | 35.7 (30.7, 41.0) | **0.34 (0.25, 0.46) \*\*** | **0.34 (0.25, 0.46) \*\*** |
| Female | 2339 (58.4) | 1475 | 58.3 (54.6, 62.0) | Ref | Ref |
| **Age (years)** |  |  |  |  |  |
| 18–29 | 1295 (32.4) | 495 | 36.8 (32.7, 41.1) | Ref | Ref |
| 30–39 | 1104 (27.6) | 576 | 46.8 (41.3, 52.4) | **1.51 (1.21, 1.90) \*\*** | **1.53 (1.19, 1.97) \*** |
| 40–49 | 736 (18.4) | 458 | 58.5 (52.4, 64.5) | **2.43 (1.94, 3.04) \*\*** | **2.74 (2.11, 3.47) \*\*** |
| 50–69 | 868 (21.7) | 579 | 63.4 (57.8, 68.7) | **2.98 (2.24, 3.97) \*\*** | **3.88 (2.85, 5.28) \*\*** |
| **Marital status** |  |  |  |  |  |
| In union | 2681 (67.0) | 1493 | 51.1 (46.8, 55.4) | Ref | Ref |
| Not-in-union | 1322 (33.0) | 615 | 39.0 (34.8, 43.3) | **0.61 (0.50, 0.74) \*\*** | **0.70 (0.54, 0.91) \*** |
| **Level of education** |  |  |  |  |  |
| No formal | 1518 (37.9) | 792 | 44.8 (41.1, 48.6) | Ref | Ref |
| Primary level | 1332 (33.3) | 672 | 42.9 (38.1, 47.8) | 0.92 (0.75, 1.14) | 0.97 (0.78, 1.22) |
| Secondary and higher | 1153 (28.8) | 644 | 52.6 (46.8, 58.3) | **1.36 (1.07, 1.74)** | **1.48 (1.09, 2.03) \*** |
| **Employment status** |  |  |  |  |  |
| Employed | 788 (19.7) | 438 | 51.2 (43.1, 59.2) | Ref | Ref |
| Self-employed | 1635 (40.8) | 851 | 46.9 (42.0, 51.8) | 0.84 (0.61, 1.16) | 0.99 (0.71, 1.36) |
| Unemployed/unpaid | 1580 (39.5) | 819 | 44.0 (40.0, 48.1) | 0.75 (0.52, 1.08) | 0.78 (0.54, 1.11) |
| **Wealth status** |  |  |  |  |  |
| 1 – Poorest | 712 (17.8) | 471 | 59.7 (52.5, 66.5) | Ref | Ref |
| 2 – Poorer | 817 (20.4) | 439 | 44.2 (38.7, 49.8) | **0.53 (0.41, 0.70) \*\*** | **0.58 (0.44, 0.78) \*\*** |
| 3 – Middle | 859 (21.5) | 438 | 47.5 (43.1, 51.9) | **0.61 (0.44, 0.85) \*** | 0.72 (0.51, 1.02) |
| 4 – Richer | 875 (21.9) | 416 | 41.7 (36.6, 46.9) | **0.48 (0.33, 0.69) \*\*** | **0.60 (0.38, 0.95) \*** |
| 5 – Richest | 740 (18.5) | 344 | 38.6 (33.9, 43.6) | **0.42 (0.30, 0.60) \*\*** | **0.52 (0.34, 0.80) \*** |
| **Smoking status** |  |  |  |  |  |
| Never/past smoker | 3539 (88.4) | 1922 | 49.0 (45.2, 52.8) | Ref | Ref |
| Current smokers | 464 (11.6) | 186 | 31.4 (24.5, 39.3) | **0.47 (0.33, 0.69) \*\*** | **0.64 (0.32, 0.98) \*** |
| **Alcohol use** |  |  |  |  |  |
| Never/past drinkers | 3128 (78.1) | 1734 | 49.4 (46.1, 52.6) | Ref | Ref |
| Current drinkers | 873 (21.9) | 374 | 39.4 (32.4, 46.8) | **0.66 (0.50, 0.88) \*** | 0.93 (0.70, 1.23) |
| **Fruit/vegetable intake** |  |  |  |  |  |
| Inadequate | 3036 (75.8) | 1615 | 48.3 (44.2, 52.5) | Ref | Ref |
| Adequate | 967 (24.2) | 493 | 40.6 (35.6, 45.9) | **0.73 (0.57, 0.95) \*** | 0.80 (0.61, 1.04) |
| **Physical activity** |  |  |  |  |  |
| High | 3003 (75.0) | 1483 | 44.7 (40.9, 48.6) | Ref | Ref |
| Moderate | 593 (14.8) | 367 | 53.7 (48.6, 58.8) | **1.44 (1.14, 1.80) \*** | 1.09 (0.83, 1.43) |
| Low | 404 (10.2) | 258 | 52.5 (44.2, 60.8) | 1.37 (0.97, 1.92) | 1.29 (0.96, 1.74) |
| **Residence** |  |  |  |  |  |
| Rural | 1992 (49.8) | 986 | 43.4 (39.6, 47.4) | Ref | Ref |
| Urban | 2011 (50.2) | 1122 | 51.6 (45.3, 57.9) | **1.39 (1.03, 1.88) \*** | 1.21 (0.89, 1.63) |
| **Region** |  |  |  |  |  |
| Rift Valley | 1239 (31.0) | 662 | 46.3 (39.3, 53.4) | Ref | Ref |
| Eastern | 700 (17.5) | 387 | 47.4 (42.8, 52.1) | 1.05 (0.74, 1.47) | 0.98 (0.67, 1.44) |
| Nyanza | 539 (13.5) | 252 | 38.9 (34.2, 43.8) | 0.74 (0.52, 1.05) | 0.75 (0.50, 1.12) |
| Coast | 493 (12.3) | 254 | 45.7 (39.3, 52.4) | 0.98 (0.66, 1.45) | 1.03 (0.70, 1.53) |
| Central | 484 (12.1) | 323 | 62.9 (56.0, 69.3) | **1.97 (1.31, 2.95) \*** | 1.54 (0.94, 2.51) |
| Western | 391 (9.8) | 139 | 30.9 (24.7, 38.0) | 0.52 (0.34, 0.79) | 0.47 (0.30, 0.75) |
| North-Eastern | 98 (2.5) | 58 | 58.9 (47.6, 69.3) | 1.66 (0.97, 2.84) | 2.27 (0.94, 5.55) |
| Nairobi | 59 (1.5) | 33 | 51.1 (34.8, 67.2) | 1.22 (0.59, 2.52) | 1.05 (0.54, 2.03) |

**Bold:** Statistically significant; \* p< 0.05, \*\* p<0.001

### Prevalence of central obesity

The overall weighted prevalence of central obesity was 46.7% (95% CI: 43.2 to 50.3), with a higher prevalence among women than men (58.3% vs. 35.6%, p=0.001). The prevalence of central obesity was highest among participants who were: from the poorest households (59.7%), secondary educated (52.6%), living in urban areas (51.6%), employed (51.2%) and married (51.1%). The sex-specific prevalence of central obesity across all variables showed similar patterns to the overall prevalence. Women exhibited a higher prevalence compared to men, with prevalence rates also rising with increasing age. (Fig. 2)

**Fig 2.**
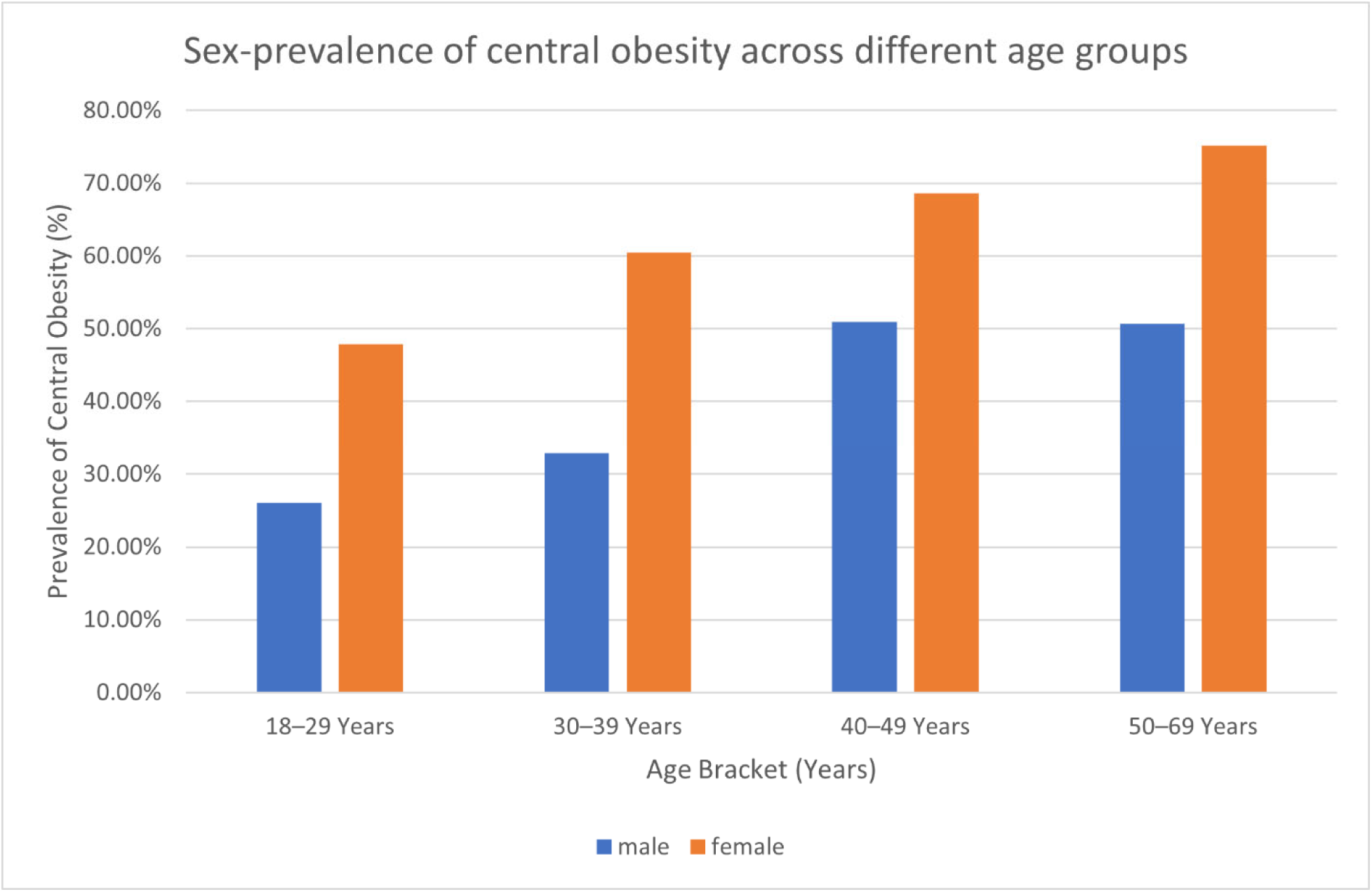
Sex-prevalence of central obesity across different age groups.

Regarding various measures of central obesity, women consistently exhibit higher rates than men, with 46.9%, 35.2%, and 46.1%, respectively, compared to 11.7%, 26.6%, and 24.2% in men.

**Table 2.** Prevalence of Central Obesity among adults by gender using different anthropometric measures.

| Measure | General |  | Men |  | Women |  |
| --- | --- | --- | --- | --- | --- | --- |
|  | n | % (95% C.I.) | n | % (95% C.I.) | n | % (95% C.I.) |
| Waist Circumference | 1,381 | 28.9 (25.9, 32.0) | 196 | 11.7 (8.3, 16.3) | 1,185 | 46.9 (43.3, 50.6) |
| Waist-to-Hip Ratio | 1,410 | 30.8(27.3, 34.4) | 467 | 26.6(21.7, 32.1) | 943 | 35.2(31.7, 38.8) |
| Waist-to-height ratio | 1,597 | 34.9 (31.7, 38.2) | 436 | 24.2(19.9, 29.0) | 1,161 | 46.1(42.3, 49.9) |

However, there were regional differences in the overall and sex-specific prevalence of central obesity. The Central (62.9%) and North-Eastern (58.9%) regions had the highest prevalence of central obesity while Western (30.9%) and Nyanza (38.9%) regions had the lowest. (Table 1).

### Factors associated with central obesity

Both in the bivariate and multivariate analysis, sex, age, marital status, level of education, and household wealth status were significantly associated with central obesity. After adjusting for all the variables, behavioural variables (smoking status, alcohol intake, vegetable and fruit intake and physical activity) were not significant in the final multivariate model. From the multivariate analysis, the odds of central obesity were 1.5- to 4-fold higher with increasing age and 1.5-fold higher among those who were secondary educated compared to 18–29-year-olds and those with no formal education, respectively (Table 1). Participants of the richest, richer, middle, and poorer wealth quantiles had 48% (aOR: 0.52, 95% CI: 0.34, 0.80), 40% (aOR: 0.60, 95% CI: 0.38, 0.95), 28% (aOR: 0.72, 95% CI: 0.51, 1.02), and 42% (aOR: 0.58, 95% CI: 0.44, 0.78) lower odds of central obesity compared to the poorest wealth quantile. Western region and Nyanza had the lowest odds of central obesity at 53% (aOR: 0.47, 95% CI: 0.30, 0.75) and 24% (aOR: 0.75, 95% CI: 0.50, 1.12) lower odds than Rift Valley. Additionally, the odds of central obesity were 66% (aOR: 0.34, 95% CI: 0.25, 0.46), 36% (aOR: 0.64, 95% CI: 0.32, 0.98), and 30% (aOR: 0.70, 95% CI: 0.54, 0.91) lower among men, smokers and unmarried compared to women, non–smokers and married, respectively (Table 1). On the sex-stratification of factors associated with central obesity, the odds of central obesity were higher among women who were older, in richer households, consumers of alcohol and those with low physical intensity as compared to men with the same characteristics (Table 3).

**Table 3.** Sex-stratified sample characteristics and prevalence and factors associated with central obesity in Kenya.

| Characteristics | Sample |  | Prevalence of central obesity |  | Multivariable logistic regression analysis |  |
| --- | --- | --- | --- | --- | --- | --- |
|  | Male | Female | Male | Female | Male | Female |
|  | n (%) | n (%) | % (95% CI) | % (95% CI) | aOR (95% CI) | aOR (95% CI) |
| **Sex** |  |  |  |  |  |  |
| Male | 1664(41.6) |  | 35.7 (30.7, 41.0) |  |  |  |
| Female |  | 2339(58.4) |  | 58.3 (54.6, 62.1) |  |  |
| **Age (years)** |  |  |  |  |  |  |
| 18–29 | 523 (31.4) | 772 (33.0) | 26.1 (20.1, 33.2) | 47.9 (42.3, 53.5) | Ref | Ref |
| 30–39 | 476 (28.6) | 628 (26.9) | 32.9 (26.5, 40.1) | 60.5 (53.8, 66.9) | 1.40 (0.87, 2.23) | **1.66 (1.15, 2.40) \*** |
| 40–49 | 320 (19.2) | 416 (17.8) | 50.9 (42.3, 59.6) | 68.6 (62.3, 74.3) | **3.02 (1.96, 4.66) \*\*** | **2.36 (1.67, 3.32) \*\*** |
| 50–69 | 345 (20.7) | 523 (22.4) | 50.7 (42.6, 58.9) | 75.2 (67.3, 81.6) | **3.93 (2.55, 6.06) \*\*** | **4.11 (2.59, 6.53) \*\*** |
| **Marital status** |  |  |  |  |  |  |
| In union | 1132 (68.0) | 1549 (66.2) | 40.8 (34.5, 47.5) | 61.3 (56.7, 65.7) | Ref | Ref |
| Not-in-union | 532 (32.0) | 790 (33.8) | 27.3 (21.2, 34.5) | 52.6 (46.7, 58.5) | 0.79 (0.52, 1.22) | **0.61 (0.43, 0.87) \*** |
| **Level of education** |  |  |  |  |  |  |
| No formal | 498 (29.9) | 1020 (43.6) | 26.3 (20.9, 32.6) | 59.3 (54.3, 64.1) | Ref | Ref |
| Primary level | 556 (33.4) | 776(33.2) | 29.2 (23.6, 35.4) | 55.5 (49.4, 61.5) | 1.25 (0.80, 1.95) | 0.81 (0.59, 1.11) |
| Secondary and higher | 610 (36.7) | 543 (23.2) | 47.4 (39.2, 55.7) | 61.0 (54.1, 67.4) | **2.11 (1.33, 3.34) \*** | 0.93 (0.56, 1.53) |
| **Employment status** |  |  |  |  |  |  |
| Employed | 490 (29.5) | 298 (12.7) | 44.1 (34.7, 54.0) | 69.0 (59.8, 76.9) | Ref | Ref |
| Self-employed | 781 (46.9) | 854 (36.5) | 36.9 (31.1, 43.2) | 59.7 (53.4, 65.7) | 1.15 (0.79, 1.68) | 0.75 (0.44, 1.26) |
| Unemployed | 393 (23.6) | 1187 (50.8) | 23.4 (17.7, 30.1) | 54.8 (50.0, 59.5) | 0.67 (0.39, 1.17) | 0.67 (0.39, 1.16) |
| **Wealth status** |  |  |  |  |  |  |
| 1 – Poorest | 348 (20.9) | 364 (15.6) | 57.8 (48.6, 66.5) | 62.5 (54.0, 70.3) | Ref | Ref |
| 2 – Poorer | 371 (22.3) | 446 (19.1) | 27.0 (20.5, 34.8) | 65.3 (58.9, 71.2) | **0.35 (0.20, 0.60) \*\*** | 1.13 (0.71, 1.83) |
| 3 – Middle | 330 (19.8) | 529 (22.6) | 32.5 (26.3, 39.3) | 59.2 (52.8, 65.2) | **0.55 (0.35, 0.89) \*** | 0.97 (0.57, 1.64) |
| 4 – Richer | 358 (21.5) | 517 (22.1) | 27.6 (20.4, 36.3) | 56.2 (50.4, 61.7) | **0.46 (0.25, 0.86) \*** | 0.79 (0.49, 1.28) |
| 5 – Richest | 257 (15.4) | 483 (20.7) | 27.0 (19.9, 35.6) | 48.3 (42.0, 54.7) | **0.48 (0.27, 0.86) \*** | **0.57 (0.33, 0.97) \*** |
| **Smoking** |  |  |  |  |  |  |
| Never/past smoker | 1288 (77.4) | 2251 (96.2) | 37.6 (31.8, 43.8) | 58.6 (54.8, 62.4) | Ref | Ref |
| Current smokers | 376 (22.6) | 88 (3.8) | 28.8 (21.4, 37.6) | 50.0 (40.2, 59.9) | 0.68 (0.39, 1.17) | 0.60 (0.34, 1.04) |
| **Alcohol use** |  |  |  |  |  |  |
| Never/past users | 981 (59.0) | 2147 (91.8) | 36.2 (30.7, 42.0) | 58.3 (54.3, 62.2) | Ref | Ref |
| Current users | 683 (41.0) | 192 (8.2) | 35.0 (28.2, 42.5) | 58.7 (46.2, 70.2) | 0.87 (0.63, 1.22) | 1.12 (0.66, 1.91) |
| **Fruit and vegetable intake** |  |  |  |  |  |  |
| Inadequate | 1236 (74.3) | 1800 (77.0) | 38.0 (32.2, 44.2) | 59.0 (54.8, 63.0) | Ref | Ref |
| Adequate | 428 (25.7) | 539 (23.0) | 27.2 (20.8, 34.8) | 55.8 (48.2, 63.2) | 0.70 (0.45, 1.08) | 0.94 (0.67, 1.33) |
| **Physical activity** |  |  |  |  |  |  |
| High | 1338 (80.4) | 1665 (71.2) | 34.0 (29.0, 39.5) | 56.5 (51.9, 61.0) | Ref | Ref |
| Moderate | 203 (12.2) | 390 (16.7) | 43.2 (31.7, 55.4) | 62.2 (53.2, 70.4) | 1.00 (0.59, 1.71) | 1.13 (0.73, 1.75) |
| Low | 123 (7.4) | 284 (12.1) | 39.6 (27.2, 53.5) | 66.3 (57.8, 73.9) | 1.07 (0.54, 2.12) | 1.36 (0.90, 2.05) |
| **Residence** |  |  |  |  |  |  |
| Rural | 771 (46.3) | 1221 (52.2) | 29.6 (25.1, 34.5) | 56.5 (51.4, 61.6) | Ref | Ref |
| Urban | 893 (53.7) | 1118 (47.8) | 43.7 (34.8, 53.1) | 61.5 (56.0, 66.6) | 1.16 (0.77, 1.76) | 1.28 (0.90, 1.83) |
| **Region** |  |  |  |  |  |  |
| Rift Valley | 537 (32.3) | 702 (30.0) | 33.1 (25.4, 41.9) | 60.1 (52.8, 67.0) | Ref | Ref |
| Eastern | 262 (15.8) | 438 (18.7) | 31.3 (24.1, 39.4) | 62.5 (56.2, 68.4) | 0.92 (0.51, 1.68) | 0.97 (0.64, 1.48) |
| Nyanza | 210 (12.6) | 329 (14.1) | 26.4 (19.9, 34.1) | 49.6 (41.2, 58.1) | 0.80 (0.43, 1.51) | 0.67 (0.40, 1.13) |
| Coast | 226 (13.6) | 267 (11.4) | 34.1 (28.7, 40.1) | 60.5 (47.0, 72.6) | 0.92 (0.56, 1.52) | 1.12 (0.65, 1.95) |
| Central | 188 (11.3) | 296 (12.7) | 54.3 (43.1, 65.1) | 71.5 (62.5, 79.1) | 1.56 (0.74, 3.26) | 1.49 (0.87, 2.58) |
| Western | 185 (11.1) | 206 (8.8) | 21.7 (16.2, 28.4) | 42.3 (29.2, 56.6) | **0.46 (0.27, 0.81) \*** | **0.45 (0.24, 0.84) \*** |
| North-Eastern | 30 (1.8) | 68 (2.9) | 54.0 (21.0, 83.8) | 62.6 (48.2, 75.1) | 3.94 (0.65, 23.94) | 1.26 (0.66, 2.42) |
| Nairobi | 26 (1.6) | 33 (1.4) | 46.9 (24.2, 70.9) | 56.3 (47.2, 65.0) | 1.34 (0.55, 3.26) | 0.72 (0.41, 1.29) |
| **Bold:** Statistically significant; * p<0.05, **p<0.001 |  |  |  |  |  |  |

## Discussion

Our study estimated the prevalence of central obesity among adults in Kenya at 46.7%, almost similar to the prevalence of central obesity in Africa at 45.7% (8). However, the prevalence was lower compared to informal urban settlements in Kenya (52.0%), semi-urban areas in Togo (48.8%), low-income population in Eastern Sudan (67.8%), and Buffalo City in South Africa (67%), which could be attributed to differences in exposure to risk factors, population size, and study design (14, 21, 40, 41). However, using a threshold of WC ≥94 cm for men and WC ≥80 cm for women, studies done among adults in Uganda, urban regions of Ethiopia, and Nigeria showed a lower prevalence of 11.8%, 37.6%, and 30.1%, respectively, as compared to our study (9, 42, 43). The high prevalence of central obesity in our study could be due to the broad definition of central obesity that we used (WHtR, WHR, or WC), thus classifying more respondents as centrally obese. Intrinsic analysis of each criterion indicated that WHtR yielded the highest weighted prevalence at 34.9%. The prevalence of CO by WC and WHR was almost similar at 28.9% and 30.8%, respectively. Furthermore, the high prevalence of central obesity in our study and as also observed in other studies might be linked to the epidemiological and demographic transition (44).

We also found women had double the prevalence of central obesity and two-thirds higher odds of central obesity compared to men; this is consistent with other studies in LMICs (8, 9, 14, 40). Similar to our findings, studies in urban settlements of Nairobi indicated that women had higher odds of central obesity than men (14). The high prevalence of central obesity among women may be attributed to lower levels of physical activity compared to men, and also, women typically have lower testosterone levels (a hormone crucial for fat breakdown in men) (45, 46). Promoting physical activity and emphasizing good nutrition among women could potentially reduce the rates of central obesity. Studies in high-income countries have shown these strategies to be effective (47, 48); however, there is a lack of evidence in the local context. Longitudinal studies in Kenya, focusing on both males and females, are needed to identify predictors and effective interventions for obesity and central obesity before definitive recommendations can be made Age was associated with central obesity, with advanced age groups having increased odds of central obesity. Similarly, other studies similar to our setting have reported a high prevalence among older participants (9, 49, 50). Younger age is a protective factor of developing central obesity due to increased metabolic activity at younger ages. In addition, as people age, fat tends to accumulate in the abdominal regions (51, 52). Promotion of exercise among older people could reduce the rates of central obesity. Evidence shows that regular physical activity, especially resistance exercise, may be more suitable as a fat-reduction strategy for older obese people (53). Regularly completing activities ranging from low-intensity walking to more vigorous sports and resistance exercises decreases the risks of developing major cardiovascular and metabolic diseases such as obesity (54).

Moreover, adults residing in the urban regions had a higher prevalence of central obesity and raised odds of 21% compared to rural residents. Similar studies in LMICs have also demonstrated a higher risk of developing central obesity among urban residents than rural residents (9, 55, 56, 57). This increased prevalence is associated with faster globalization and urban dwellers’ preferences for unhealthy diets and sedentary lifestyles (58, 59, 60). Furthermore, the higher odds of central obesity in urban areas may be attributed to the overreliance on motorized means of transport (61). Prioritization in the construction of footpaths and recreational areas in urban regions could increase physical activity levels in urban areas and, therefore, reduce the rates of central obesity (62).

Socioeconomic status (SES), as measured by level of education, employment status, and household wealth, was associated with central obesity. Participants who were secondary educated or employed had higher odds of central obesity. However, participants from the poorest households had the highest prevalence and higher odds of central obesity than those from the richest households. Our findings reflect the inconclusive nature of the association between socioeconomic status and central obesity, with some studies done in an almost similar setting to ours showing a positive association between high SES and central obesity (8, 55, 63). In contrast, others show a negative association (20, 64, 65). The observed wealth inequality in central obesity in our study could be explained by limited access to healthy food such as fruits and vegetables resulting in the consumption of non-nutritious, calorie-dense foods (66). Moreover, among the reasons for the increased prevalence of obesity in the population of poor people are high levels of physical inactivity which is associated with a lack of money for sports equipment(67).

Respondents in a union (married) had raised odds of being centrally obese compared to those who were not in a union. Similarly, other studies done in Uganda (9), South Africa (21), and Ethiopia (57) established that the odds of central obesity were higher among married or cohabiting participants than among those who had never married before. The plausible reasons for higher levels of obesity among married participants could be the less emphasis on being physically attractive and dietary changes in marriage. Also, married individuals have solid social support that ensures adequate food security (68).

Smoking and alcohol use were negatively associated with central obesity, showing a protective effect, with non- or past-users having lower odds of central obesity. However, women who were using alcohol had 12% higher odds of central obesity compared to non-users of alcohol. Systematic reviews and meta-analyses have shown varying positive and negative associations between alcohol consumption and obesity among men and women (69, 70). Alcoholic drinks are calorie-dense (71) and impede fat oxidation through the antilipolytic properties of alcohol metabolites (72), which promotes fat accumulation and elevates the risk of developing adiposity (73).

Our findings show that physical inactivity is associated with higher odds of central obesity, similar to previous studies in India (74), Ghana (75), and Nigeria (76). The prevalence of central obesity was highest among respondents with moderate levels of physical intensity. Physical activity increases the amount of energy used by the body (77), while inactivity causes an imbalance with extra energy stored as fat in the abdominal area, contributing to central obesity (78, 79). It is, therefore, recommended that health education on the importance of engaging in moderate-to-high-intensity physical activity be intensified for the prevention of central obesity (58). In addition, an environment with footpaths, bicycle lanes, and public parks has been shown to encourage physical activities (62).

We observed regional differences in the prevalence of central obesity in Kenya. Over half of the respondents had central obesity in Central, North-Eastern, and Nairobi regions. The reasons for the high prevalence of central obesity in these regions are unknown. However, the high prevalence could be associated with the diet in these regions, with the Central region participants mainly consuming potatoes and a high sugar diet and participants of the North-Eastern regions having low physical activity levels due to high temperatures. Also, these regions have been shown to have a high prevalence of non-communicable diseases (80). Therefore, intensified and targeted interventions to address central obesity should be implemented in these regions, especially the marginalized North Eastern region.

### Strengths and limitations

We used the 2015 Kenya stepwise survey dataset, which is a representative large sample size. Additionally, the study included all the measures (W.C, WHR, WHtR) used to define central obesity, according to the WHO. Nonetheless, this study had certain limitations. The cross-sectional design of the survey restricted our ability to draw causal inferences. Additionally, this study majored in the determinants at the national level rather than at the county level. Therefore, future research should aim to explore the pattern of central obesity and determinants at the county level to inform more targeted policy interventions. Also, health behavioural variables such as smoking, alcohol intake, fruit and vegetable intake and physical activity relied on self-reporting which may lead to potential under or over-reporting bias. Lastly, comparing the findings to other studies may differ depending on the central obesity measure used. While some studies used only one measure of central obesity this may establish a lower prevalence than our study, which incorporated all the measures.

## Conclusion

In conclusion, almost half of Kenyan adults are centrally obese, with the prevalence being twice as high among women as compared to men. The established high rates of central obesity in Kenya warrant community sensitization. In addition, the government should formulate policies aimed at early detection, continuous monitoring, and reversal of central obesity. Future studies should venture into normal-weight central obesity to picture the interplay between general obesity and central obesity.

## Data Availability

Data used in this study can be accessed from the Kenya National Bureau of Statistics website at http://statistics.knbs.or.ke/nada/index.php/catalog.

## List of abbreviations

BMI: Body Mass Index
CO: Central Obesity
CI: Confidence Interval
LMICs: Low–and Middle–Income Countries
NCDs: Non-communicable Diseases
OR: Odds Ratio
WC: Waist Circumference
WHR: Waist-to-Hip Ratio
WHtR: Waist-to-Height Ratio
WHO: World Health Organization

## Declaration

## Ethics approval and consent to participate

We utilized secondary data from the 2015 Kenya STEPwise Survey for Non-communicable Diseases Survey approved by the Kenya Medical Research Institute Scientific and Research Ethics Units.

## Consent for publication

Not applicable

## Author contributions

SMG conceptualized, acquired, and analysed data for the study. CN, SK, and RWN conducted the literature review, interpreted the findings, and wrote the first draft. SMG provided the overall project supervision. All the authors critically reviewed, revised, read, and approved the manuscript.

## Acknowledgment

We extend our gratitude to the Kenya National Bureau of Statistics for granting us access to the data from the Kenya Stepwise Survey on non-communicable diseases.

