## Supplemental Table 1 for "Prevalence and determinants of central obesity among adults 18–69 years in Kenya: a cross-sectional study"

### Supplementary Table 1. Operational definition of variables

| **Variables** | **Operational Definition** |
| --- | --- |
| Regions | The 47 counties were clustered into eight regions: Nairobi, Central, Western, North-Eastern, Western, Nyanza, Rift Valley, and Coast. |
| Residence | Rural or urban |
| Age (years) | Age was categorized into 18–29, 30–39, 40–49, 50–69 years |
| Sex | Sex was recoded as either male or female based on the interviewer’s observations |
| Marital Status | Individuals who were married or cohabiting were grouped as in union, while single, separated, widowed, or divorced individuals were considered not in union. |
| Education level | Categorized into no formal education, primary education, and secondary and higher. |
| Employment | Employed or unemployed |
| Household wealth index | Five wealth quintiles (poorest, poorer, middle, richer, richest) were computed based on the wealth index generated using principal component analysis of the household characteristics such as the type of house, floors, ownership of durable assets such as cars and land and, access to basic services such as tap water & electricity. |
| Smoking | A different form of tobacco use was assessed and recoded as either past smoker/non-smoker or current use of smokeless tobacco and other tobacco products. |
| Alcohol use | Alcohol consumption was measured based on current and never/past use of drinks that contain alcohol. |
| Fruits and vegetable intake | Inadequate or adequate consumption was defined as an intake of less than five servings of fruits and vegetables in a week or more than five servings, respectively. |
| Physical Activity | The intensity of physical activity was assessed using the Global Physical Activity Questionnaire (GPAQ). It measures the intensity, duration, and frequency of physical activity in work-, leisure- and transport-related activities. The total time spent, number of days, and intensity of physical activity in a typical week were used to generate three levels of physical activity: low, moderate, and high. |
